# Excess deaths in people with cardiovascular diseases during the COVID-19 pandemic

**DOI:** 10.1101/2020.06.10.20127175

**Authors:** Amitava Banerjee, Suliang Chen, Laura Pasea, Alvina G. Lai, Michail Katsoulis, Spiros Denaxas, Vahe Nafilyan, Bryan Williams, Wai Keong Wong, Ameet Bakhai, Kamlesh Khunti, Deenan Pillay, Mahdad Noursadeghi, Honghan Wu, Nilesh Pareek, Daniel Bromage, Theresa A. McDonagh, Jonathan Byrne, James T.H. Teo, Ajay M Shah, Ben Humberstone, Liang V. Tang, Anoop S. V. Shah, Andrea Rubboli, Yutao Guo, Yu Hu, Cathie L. M. Sudlow, Gregory Y. H. Lip, Harry Hemingway

## Abstract

**Background:** Cardiovascular diseases(CVD) increase mortality risk from coronavirus infection(COVID-19), but there are concerns that the pandemic has affected supply and demand of acute cardiovascular care. We estimated excess mortality in specific CVDs, both “direct”, through infection, and “indirect”, through changes in healthcare.

**Methods:** We used population-based electronic health records from 3,862,012 individuals in England to estimate pre- and post-COVID-19 mortality risk(“direct” effect) for people with incident and prevalent CVD. We incorporated: (i)pre-COVID-19 risk by age, sex and comorbidities, (ii)estimated population COVID-19 prevalence, and (iii)estimated relative impact of COVID-19 on mortality(relative risk, RR: 1.5, 2.0 and 3.0). For “indirect” effects, we analysed weekly mortality and emergency department data for England/Wales and monthly hospital data from England(n=2), China(n=5) and Italy(n=1) for CVD referral, diagnosis and treatment until 1 May 2020.

**Findings:** CVD service activity decreased by 60-100% compared with pre-pandemic levels in eight hospitals across China, Italy and England during the pandemic. In China, activity remained below pre-COVID-19 levels for 2-3 months even after easing lockdown, and is still reduced in Italy and England. Mortality data suggest indirect effects on CVD will be delayed rather than contemporaneous(peak RR 1.4). For total CVD(incident and prevalent), at 10% population COVID-19 rate, we estimated direct impact of 31,205 and 62,410 excess deaths in England at RR 1.5 and 2.0 respectively, and indirect effect of 49932 to 99865 excess deaths.

**Interpretation:** Supply and demand for CVD services have dramatically reduced across countries with potential for substantial, but avoidable, excess mortality during and after the COVID-19 pandemic.

**Funding:** NIHR, HDR UK, Astra Zeneca

## Introduction

Coronavirus disease 2019(COVID-19) has had unplanned consequences for non-COVID-19 health services. We described reductions in urgent cancer referrals and chemotherapy compared to pre-COVID-19 levels^1^. Decreases in presentation and treatment of myocardial infarction(MI)^2–4^ in Italy and the USA suggest effects on care, but services have not been studied across specific CVDs, countries or different phases of the pandemic.

Early reports from Wuhan, China, demonstrated high prevalence of and mortality from COVID-19 in individuals with CVD; confirmed across countries, particularly with coronary heart disease(CHD) and heart failure(HF)^5–8^. UK government policy for “physical distancing” in high-risk subgroups for COVID-19, announced on 16 March 2020, included CVD, especially HF^9^. On 22 March 2020, a further 1.5 million people in England(with ‘extremely vulnerable’ conditions) were recommended at least 12 weeks of ‘shielding’^10^, excluding those with CVD, prior to UK lockdown on 23 March^11^. Better understanding of pre- and post-COVID-19 mortality risk across specific CVDs may help decisions and policies regarding physical isolation.

Therefore, effects of COVID-19 on individuals and health systems are: (i)*direct* due to infection, and (ii)*indirect* due to unprecedented system strain and associated behaviour changes^12^. Those with CVD, carrying the greatest burden of global morbidity and mortality^13^, are likely to be particularly affected. Beyond direct effects, systematic examination of recent and longer-term trends for CVD services may help in planning the timing and nature of exit from lockdown and strategies for any subsequent infection peaks. Moreover, reductions in rates of referral, diagnosis, and treatment, may have fatal long-term consequences, caused by “supply”(e.g. lower healthcare availability) and/or “demand”(e.g. delayed presentation), but have not been investigated. Electronic health records(EHR) have been used in studies across different specific CVDs^14^, and enable novel insights regarding direct and indirect COVID-19-related excess deaths, across incident and prevalent CVD, and the spectrum of care.

### Objective

We used: (i)national mortality data for England and Wales to investigate trends in non-COVID-19 and CVD excess deaths; (ii)routine data from hospitals in England, Italy and China to assess indirect effects on services for referral, diagnosis and treatment of CVD during the pandemic; and (iii)population-based EHR in England to investigate pre-COVID-19 mortality by underlying risk factors and excess mortality during the COVID-19 pandemic.

## Methods

### Data sources and study population

To estimate pre-COVID-19 incidence and mortality in individuals with CVD and comorbidities, we used EHR across primary care(Clinical Practice Research Datalink, CPRD-GOLD), hospital care(Hospital Episodes Statistics, HES), and death registry(Office of National Statistics, ONS) with prospective recording and follow-up; linked by CPRD and NHS Digital using unique healthcare identifiers^15^. Over 99% of England’s population is registered with general practice(GP). CPRD is representative by socio-demography, ethnicity and overall mortality^16^. Eligible individuals were aged ≥30 years, registered with a GP between 1 January 1997 and 1 January 2017 with ≥1 year of follow-up. Study entry was 1 year following latest GP registration for each individual, to more completely capture past medical history.

### Open-access electronic health record phenotypes

We defined non-fatal(alive for ≥30 days post-diagnosis) CVD using 16 previously validated CALIBER phenotypes: stable angina, unstable angina, MI, CHD unspecified, HF, cardiac arrest, transient ischaemic attack(TIA), ischaemic stroke, stroke unspecified, intracerebral haemorrhage, subarachnoid haemorrhage, peripheral arterial disease(PAD), atrial fibrillation(AF), abdominal aortic aneurysm(AAA), DVT and pulmonary embolism(PE), as per prior studies^14–16^. We included DVT and PE due to clinical importance of venous thromboembolism in COVID-19^17^. Incident and prevalent CVD were defined by first CVD record in the study period with no prior CVD history, and first prior CVD history, respectively. Validated phenotype definitions of diseases and COVID-19-relevant conditions(available at https://caliberresearch.org/portal) ^14–16^ were generated from hospital and primary care information recorded in primary care, using Read clinical terminology(version 2).

We defined 15 comorbidities or comorbidity clusters involving 40 individual conditions associated with poor COVID-19 outcomes by UK government guidance^9,10^. Further details are provided in **Supplementary Methods**. Multimorbidity was co-occurrence of ≥2 of these conditions^18^.

### Weekly/monthly information on CVD care and mortality

We obtained: (i)weekly cause-specific mortality data for England and Wales from ONS^19^ and cardiac-related attendance data from the Emergency Department(ED) Syndromic Surveillance System for England^20^; (ii)monthly referral, diagnosis and treatment data for CVD in two UK hospitals(University College London NHS Trust, and King’s College Hospital London NHS Foundation Trust); and (iii)hospital data in China(n=5; Wuhan Union Hospital, Zaoyang First Hospital, Xiangyang First Hospital, Xiantao People’s Hospital, Tianmen First Hospital) and Italy(n=1; Ospedale Santa Maria delle Croci, Ravenna). For all data, weekly or monthly relative risks(RRs) were calculated compared with the pre-lockdown data(from 3 January). Further details of data and RR calculations are given in the **Supplementary Methods**.

### Estimating incidence rates and 1-year mortality

We estimated incidence rates per 100,000 person-years and pre-COVID-19 1-year mortality risk for incident and prevalent CVD using Kaplan-Meier analyses stratified by specific CVD and number of (non-CVD) comorbidities(0, 1, 2 and 3+), scaling up from CALIBER(3.8 million individuals) to the whole population of England aged 30+, consisting of 35,407,313 individuals(using 2018 estimates of overall population size and mortality^21^).

### Estimating 1-year direct and indirect excess deaths

Excess deaths were considered as direct(due or with COVID-19 infection) or indirect(due to changes in health services). *Direct excess deaths* were estimated by applying RRs of 1.2, 1.5, 2 and 3, based on published hazard ratios for CVD and COVID-19 deaths^8,22,23^, in the absence of cohort studies of clinical cohorts of CVD patients investigating all-cause mortality in those with and without infection. We modelled 10% infection rate based on recent COVID-19 seroprevalence estimates^24–25^. Although infection rate will change depending on pandemic phase, we assumed infection rate over 1 year in line with the first wave.

Given service changes in CVD observed across countries^2–4^, to capture *direct* and *indirect* excess deaths, we modelled in addition to the 10% infected, 40% and 80% affected rates, where 30% and 70% of the population respectively would be un-infected but affected at corresponding RRs. Based on RRs from ONS data(peak RR 1.14 for excess CVD deaths and 1.29 for excess non-COVID-19 deaths; **Figure 2**) and likely longer-term effects on CVD mortality, we estimated direct and indirect excess deaths together by applying RR 1.2 to 40% and 80% of the population. Thus, we provide *low*(infection rate 10% and no indirect effect at RR 1.5, 2.0 and 3.0), *medium*(infection rate 10% and 30% indirectly affected at RR 1.2) and *high* estimates(infection rate 10% and 70% indirectly affected at RR 1.2) of excess deaths. We projected excess death estimates to the whole English population, we used the 2018 population estimate of 35,407,313 individuals aged ≥30^21^. All analyses were performed using R(version 3.4.3).

## Results

### Monthly hospital data across countries

In China (both Wuhan and other hospitals) and Italy, activity(referral, diagnosis and treatment) was significantly reduced compared with pre-COVID levels. In Wuhan, there was 83.4% reduction in referral, 92% in diagnosis and 99% in treatment levels of MI by the 1 February, a month after the outbreak started. Corresponding decreases were 92%, 97% and 87% for stroke, 83%, 93% and 98% for HF, and 83%, 95% and 94% for AF. Tianmen and Xiantao demonstrated equivalent decreases across all four disease services. Although decreases were relatively less in Zaoyang and Xiangtang, there were 40-80% reductions across services, except for diagnosis of MI which increased in Zaoyang between January and March.

Even after easing and stopping of lockdown, Wuhan reported 50-100% decreases in services for MI, stroke, HF and AF. In other Chinese hospitals, activity had mostly returned to pre-COVID levels by April 2020. Recovery of activity began to occur during the lockdown period and differed across Chinese hospitals, suggesting possible differences in infection rates, patient presentation and behaviour, clinician behaviour and COVID-19-related system strain. In Italy, decreases ranged from 45% for stroke treatment to 94% for HF treatment and 100% for AF diagnosis. In England, there was reduced activity across CVDs during the pandemic. Services in Italy and England had not reached pre-COVID levels even by the easing of lockdown. Across countries, referral rates for all diseases and treatment of MI were particularly affected and declines in activity occurred before the peak of cases or deaths(**Figure 1**).

**Figure 1:**
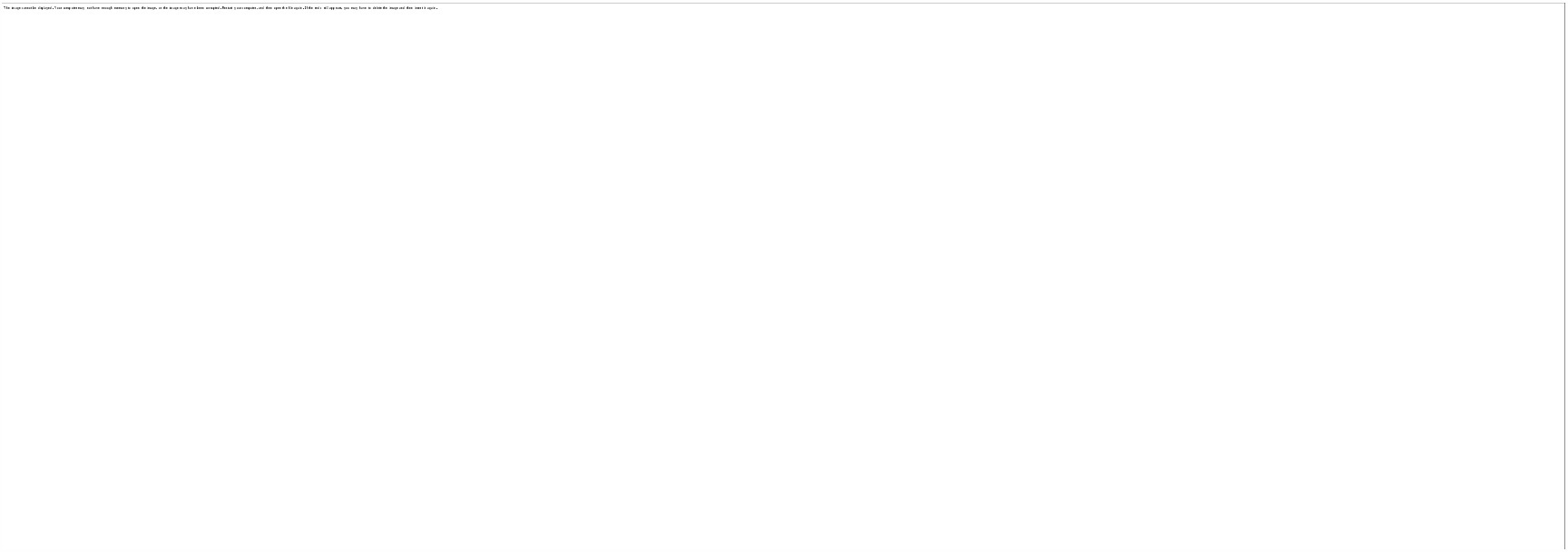
Hospital activity before and during the COVID-19 pandemic for referral, diagnosis and treatment of cardiovascular disease in China, Italy and England.

### Weekly national mortality data and emergency attendance data

Non-COVID-19 and CVD-related deaths in England and Wales increased in the same chronology as total(including COVID-19) deaths until mid-May 2020(compared with the same week over the last 5 years). The peak observed RR for CVD deaths was in the week ending 24 April(1850 vs 1626 deaths, RR 1.14) and the lowest in the week ending 8 May 2020(1318 vs 1487 deaths, RR 0.89). The same trend was present regardless of whether the RR was calculated compared to the average of previous years, or pre-COVID-19(3 January 2020). Between 6 and 27 March, cardiac ED attendances decreased in England(minimum RR 0.57) and were had not fully recovered on 15 May, which was after easing of lockdown(**Figure 2**).

**Figure 2.**
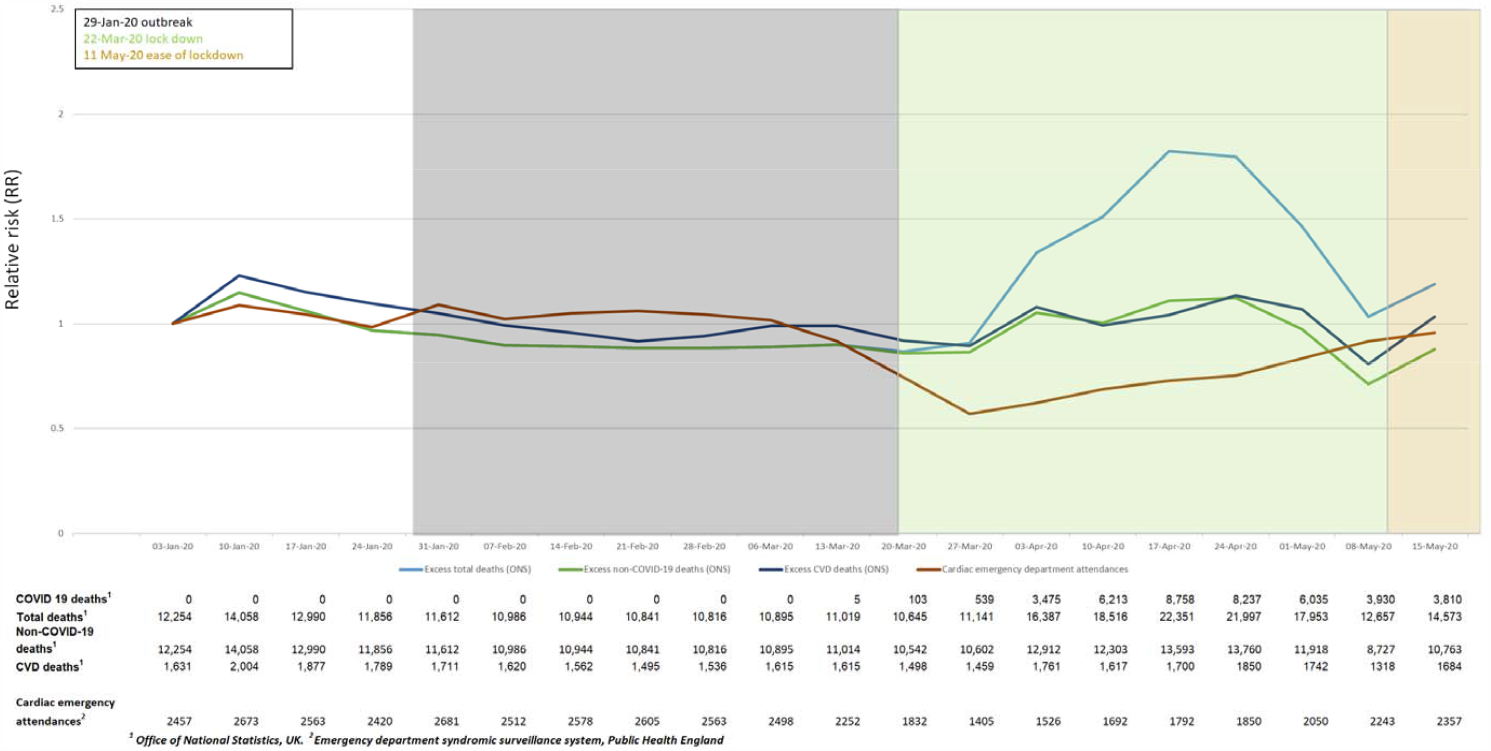
Weekly national data for excess total, non-COVID and cardiovascular deaths in England and Wales, and emergency department cardiac attendances for England: relative risks.

### Incidence, prevalence and pre-COVID-19 mortality for CVD

Of 3,862,012 individuals in our English cohort, 538,037 had incident CVD and 580,437 had prevalent CVD. Age-adjusted incidence rates(per 100,000 population) were highest for AF(330), CHD(261), HF(168), stable angina(140), and DVT(139)(**Figure S1**). For incident CVDs, 1-mortality rates varied by specific CVD: 58.4% for cardiac arrest, 42.1% for intracerebral haemorrhage, 30.3% for HF, 20.1% for MI, 12.1% for CHD, 4.8% for stable angina,. Absolute 1-year mortality was highest for AF(234,778), HF(173,955), CHD(114,383) and stroke(63,276). Mortality rates were relatively lower for prevalent CVDs(**Figure S2**).

### Comorbidities in individuals with CVD

Across incident and prevalent CVDs, prevalence of 0, 1, 2 and 3+ co-morbidities was 26.2% and 27.1%, 33.2% and 36.3%, 21.9% and 20.5%, 18.6% and 16.2% respectively. Across CVDs, hypertension, CKD, cancer, chronic obstructive pulmonary disease(COPD) and diabetes were commonest(**Supplementary Table 1, Figure S3**). Comorbidity profiles were similar in those with prevalent CVD(**Figure S4**). Multimorbidity was associated with increased 1-year mortality, e.g. in incident AF, 1-year mortality was 31% vs 9% in men and 34% vs 16% in women for ≥3 vs 0 conditions. One-year mortality risks were >50% in those with incident cardiac arrest or intracerebral haemorrhage and ≥3 conditions(**Figures 3B** and **S5B**).

**Figure 3:**
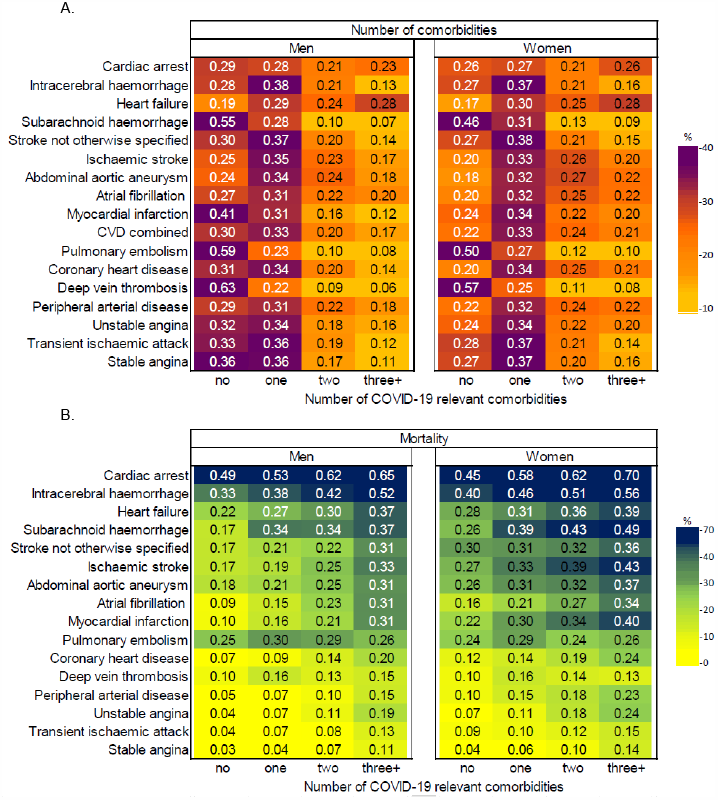
COVID-19 relevant comorbidities in incident cardiovascular diseases in England: (A) Proportion of individuals with 0, 1, 2 and 3+ comorbidities by specific CVD. (B) 1-year mortality in individuals with 0, 1, 2 and 3+ comorbidities by specific CVD.

### Estimates for direct and indirect excess 1-year COVID-19-related deaths

For incident CVD, at 10% infection rate, there would be 5067, 10,135 and 20,269 excess deaths at RR 1.5, 2.0 and 3.0 respectively. At 40% and 80% infection rates, we estimated 8112 and 16225 excess deaths at RR 1.2 (**Figure 4A**).

**Figure 4:**
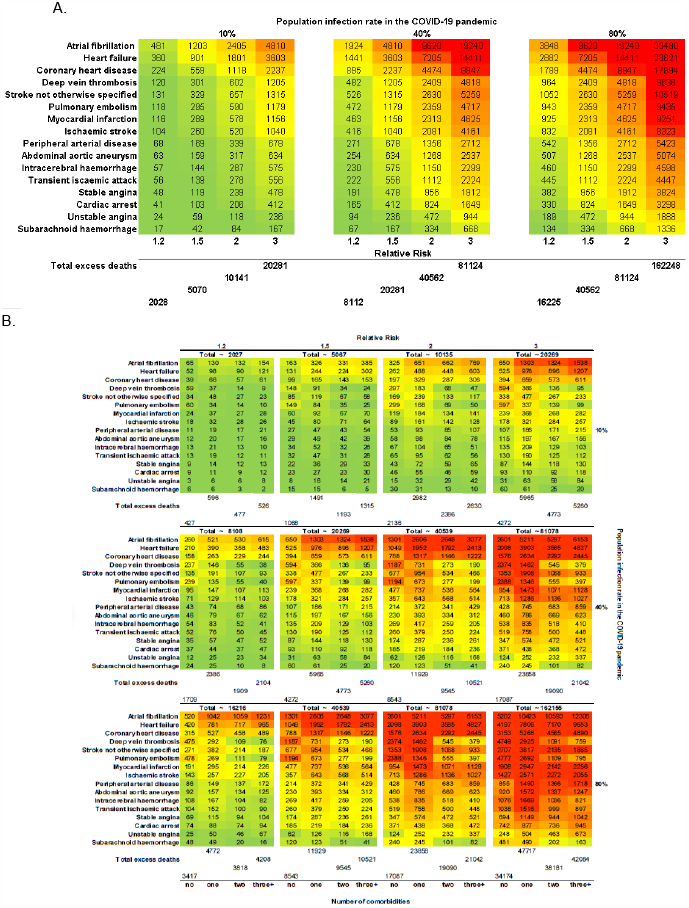
Estimated number excess deaths at 1 year due to COVID-19 pandemic by CVD type for: (A) incident cases; and (B) number of comorbidities for incident cases.

For prevalent CVD, at 10% infection rate, there would be 26,138, 52,275 and 104,550 excess deaths at RR 1.5, 2.0 and 3.0 respectively. At 40% and 80% infection rates, we estimated 41820 and 83640 excess deaths at RR 1.2(**Figure 5A**).

**Figure 5:**
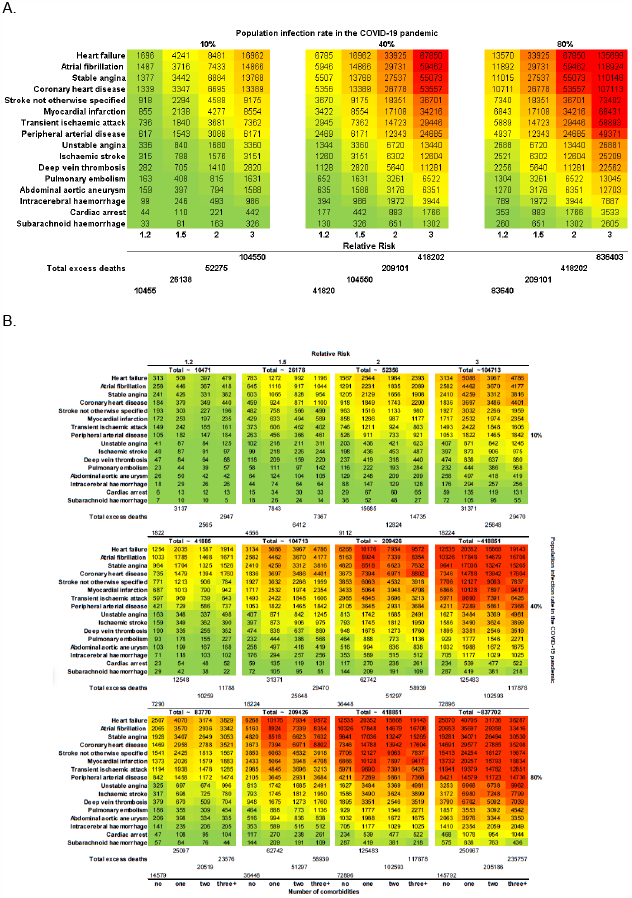
Estimated number excess deaths at 1 year due to COVID-19 pandemic by CVD type for: (A) prevalent cases; and (B) number of comorbidities for prevalent cases

For total CVD(incident and prevalent combined), at 10% infection rate, we estimated a direct effect of 31,205, 62,410, and 124, 819 excess deaths at RR, 1.5, 2.0 and 3.0 respectively. For total CVD, we estimate an indirect effect of 49932 to 99865 excess deaths using 40% and 80% infection rate estimates.

For incident CVD, at 10% COVID-19 rate, we estimated 2508 vs 1068, 5016 vs 2136 and 10033 vs 4272 excess deaths in multimorbidity compared with no comorbidities at RR 1.5, 2.0 and 3.0 respectively. At 40% COVID-19 rate, we estimated 4013 vs 1709 excess deaths; and at 80% COVID-19 rate, we estimated 8026 vs 3417 excess deaths in multimorbidity compared with no comorbidities at corresponding RRs(**Figure 4B**). Among those with prevalent CVDs, we estimated higher excess deaths at 1 year than for incident CVDs with similar associations with number of comorbidities(**Figure 5B**).

## Discussion

This first analysis from large-scale population-based EHR across 16 specific CVDs and multimorbidity in the COVID-19 context has four findings. First, profound disruption of CVD services across referral, diagnosis and treatment(seen in UK, Italy and China) may contribute to excess deaths. Second, there are high rates of “high-risk” and “extremely vulnerable” conditions among people with CVD, often in combination, varying by specific CVD. Third, we predict significant excess deaths in individuals with CVD, over a 1-year time horizon partly because indirect effects may be delayed. Fourth, excess deaths in prevalent and incident CVD, by direct and indirect effects, suggest that access to acute and chronic CVD care should be prioritised during future pandemic waves.

Professional organisations quickly produced evidence-based CVD management guidelines in the COVID-19 context^26^. Our analysis highlights the pandemic’s real and potential impact on CVD healthcare provision in different countries at different stages in their responses. At peak, almost all CVD healthcare activity collapsed in Wuhan. Although Italy and the UK were affected later, CVD services were, and still are, compromised in pre-lockdown and lockdown phases. Changes in CVD excess deaths have not yet been observed in ONS data, suggesting that indirect effects may manifest over at least a year, rather than contemporaneously with activity reductions observed across countries. Overall, these data emphasise indirect effects of COVID-19, helping to quantify and model true “relative impact”, and relative contributions of pandemic versus lockdown on CVD services. We believe countries should monitor near real-time service activity, and COVID-19 and non-COVID-19 deaths^27^ over the coming year to understand and mitigate excess deaths in individuals with CVD, especially in those countries still at earlier phases, such as Brazil.

There is debate about which conditions should be on “high-risk” and “extremely vulnerable” lists of conditions for physical isolation policies, which should include some CVDs and combinations of comorbidities, based on our risk estimates and COVID-death data to-date^6,12^. CVD is known to have high prevalence, incidence and mortality, varying by arterial territory. We now show that AF, CHD, HF, stable angina, and DVT have the highest pre-COVID-19 incidence rates. Prognosis varies by incident versus prevalent disease, specific CVD and number and combination of comorbidities, highlighting need for individualised risk prediction across CVD. On a different note, our findings also bring into question the rationale for looking at composite endpoints such as “MACE”(major adverse cardiovascular events) in trials and other studies due to highly variable risk across specific CVD^28^.

In individuals with CVD at 10% population infection rate, there would be 50,937 excess deaths at RR 1.5, with higher rates at higher infection rates. An earlier lockdown, as in New Zealand or the Indian state of Kerala, minimises overall deaths. Conversely, in countries such as the UK and USA, delayed lockdown may exacerbate direct and indirect effects of COVID-19, supported by rising non-COVID-19 deaths in ONS data. COVID-19 and non-COVID-19 deaths can be avoided by: (i)acting early and reducing infection rates by widespread testing, stringent physical isolation and suppression policies, and (ii) focusing on the preventable CVD burden. For example, HF, MI, ischaemic stroke and AF are prevalent conditions with highest mortality rates; all with evidence-based therapies. The same is true for common CVD comorbidities, which often occur in clusters: hypertension, CKD, cancer, COPD and diabetes, underlining importance of integrated CVD and risk factor management pre- and post-COVID-19.

Our findings may have implications for which aspects of health services(acute vs chronic, treatment vs prevention, across specific CVDs) require attention at different pandemic phases. Demand for NCD care is documented in humanitarian emergencies^29^, but NCD surveillance is absent in pandemic preparedness, planning and responses, particularly in low-middle income countries^13^, where our findings will be magnified. The learning health system concept^30^, where near real-time data inform science, evidence and care, has not been used optimally during the COVID-19 pandemic, but our CVD referral, diagnosis and treatment data show that data need not be complex.

## Strengths and limitations

Our study uses large-scale, nationally representative EHR with validated definitions across a comprehensive list of specific CVDs and comorbidities. We present real-time service data from three countries. There are several limitations. In the UK, our study population was 5% of the overall population. We do not have country-level data outside the UK. Our analyses of risk and excess deaths use retrospective EHR data. Our model assumes static infection rate and RR and do not include changes over time. We used service data from limited hospitals and only UK(ONS) administrative data was available. We do not report primary care or community level data regarding CVD services. We modelled using limited comorbidities and simple multimorbidity counts, and did not study impact of ethnicity. We assumed the same effect across all specific CVDs.

## Conclusion

There is a substantial avoidable burden of excess mortality in people with CVD during the COVID-19 pandemic. Excess CVD deaths can be reduced in more integrated approaches, focused on reducing COVID-19 infection rates, as well as managing CVD and comorbidities.

## Data Availability

There are no data available in relation to this manuscript.

## Contributor and guarantor information

Research question: AB, GYHL, HH Funding: AB, HH

Study design and analysis plan: AB, SC, AL, LP, HH

Preparation of data, including electronic health record phenotyping in the CALIBER open portal: SC, LP, SD

Provision of monthly hospital data: JT, AS, DB, TM, NP, JM, WKW, BW, AR, LVT, YG, YT, GYHL.

Provision of weekly cause-specific mortality data: VN, BH Provision of EDSS data: MK

Statistical analysis: SC, LP, AB

Drafting initial versions of manuscript: AB

Drafting final versions of manuscript: AB, GYHL, HH

Critical review of early and final versions of manuscript: All authors

The corresponding author attests that all listed authors meet authorship criteria and that no others meeting the criteria have been omitted. AB and HH are the guarantors.

### Ethical approval

This study was approved by the Independent Scientific Advisory Committee of the CPRD(20_074R) under Section 251(NHS Social Care Act 2006) of the UK Medicines and Healthcare products Regulatory Agency. Hospital data was obtained with approval of the relevant hospital institutional review boards.

## Funding acknowledgements

AB has received research funding from NIHR, UKRI, British Medical Association and Astra Zeneca. BW and HH are funded by the National Institute for Health Research (NIHR) University College London Hospitals Biomedical Research Centre. HH is supported by Health Data Research UK (HDR-UK; grant no LOND1), which is funded by the UK Medical Research Council, Engineering and Physical Sciences Research Council, Economic and Social Research Council, Department of Health and Social Care (England), Chief Scientist Office of the Scottish Government Health and Social Care Directorates, Health and Social Care Research and Development Division (Welsh Government), Public Health Agency (Northern Ireland), British Heart Foundation, and Wellcome Trust. AB, SC, SD, HH is also supported by the BigData@Heart Consortium, funded by the Innovative Medicines Initiative-2 joint undertaking under grant agreement no 116074. This joint undertaking receives support from the EU’s Horizon 2020 research and innovation programme and European Federation of Pharmaceutical Industries and Associations. AMS is supported by the British Heart Foundation (CH/1999001/11735), the NIHR Biomedical Research Centre at Guy’s & St Thomas’ NHS Foundation Trust and King’s College London (IS-BRC-1215-20006), and the Fondation Leducq. KK is supported by NIHR ARC East Midlands. BW is supported by the NIHR University College London Hospitals Biomedical Research Centre. HW is supported by Medical Research Council and Health Data Research UK Grant (MR/S004149/1), Industrial Strategy Challenge Grant (MC_PC_18029) and Wellcome Institutional Translation Partnership Award (PIII054). MN is supported by a Wellcome Trust Investigator in Science award (207511/Z/17/Z). DB is supported by the NIHR (CL-2016-17-001).

### Research in Context

#### Evidence before this study

Coronavirus disease 2019 (COVID-19) can cause excess deaths through direct due to infection) and indirect effects. We searched PubMed, medRxiv, bioRxiv, arXiv, and Wellcome Open Research for peer-reviewed articles, preprints, and research reports on mortality and cardiovascular disease (CVD) in (COVID-19), using the search terms “coronavirus”, “COVID-19”, “cardiovascular disease” and similar terms, and “mortality”, up to May 31, 2020. We found no prior studies of excess deaths in those with cardiovascular disease and the COVID-19 pandemic. Health services for myocardial infarction appear to have decreased in some countries, but services for CVD have not been studied across specific CVDs, countries or different phases of the pandemic. Such data will enable better prediction of the direct and indirect effects of COVID-19 and planning for health system responses.

#### Added value of this study

Comparing hospital data from the UK, Italy and China, we show a profound disruption of CVD services across referral, diagnosis and treatment which may contribute to excess deaths. Using a published model, we estimated the excess COVID-19-related mortality in individuals with CVD by incorporating population infection rate in different scenarios, differing degrees of impact of COVID-19 on health systems, and prevalence of underlying conditions. As well as high rates of multimorbidity, we predict high excess deaths in individuals with CVD, over a 1-year time horizon partly because indirect effects may be delayed.

#### Implications of all the available evidence

Our findings regarding excess deaths in prevalent and incident CVD, by direct and indirect effects, suggest that access to acute and chronic CVD care should be prioritised during future pandemic waves, as well as efforts and strategies to minimise infection rate in the population.

## Figures and Tables

Figure 1. Hospital activity before and during the COVID-19 pandemic for referral, diagnosis and treatment of cardiovascular disease in China, Italy and England.

Figure 2. Weekly national data for excess total, non-COVID and cardiovascular deaths in England and Wales, and emergency department cardiac attendances for England.

Figure 3: COVID-19 relevant comorbidities in incident cardiovascular diseases in England: (A) Proportion of individuals with 0, 1, 2 and 3+ comorbidities by specific CVD. (B) 1-year mortality in individuals with 0, 1, 2 and 3+ comorbidities by specific CVD.

Figure 4: Estimated number excess deaths at 1 year due to COVID-19 pandemic by CVD type for: (A) incident cases; and (B) number of comorbidities for incident cases

Figure 5: Estimated number excess deaths at 1 year due to COVID-19 pandemic by CVD type for: (A) prevalent cases; and (B) number of comorbidities for prevalent cases.

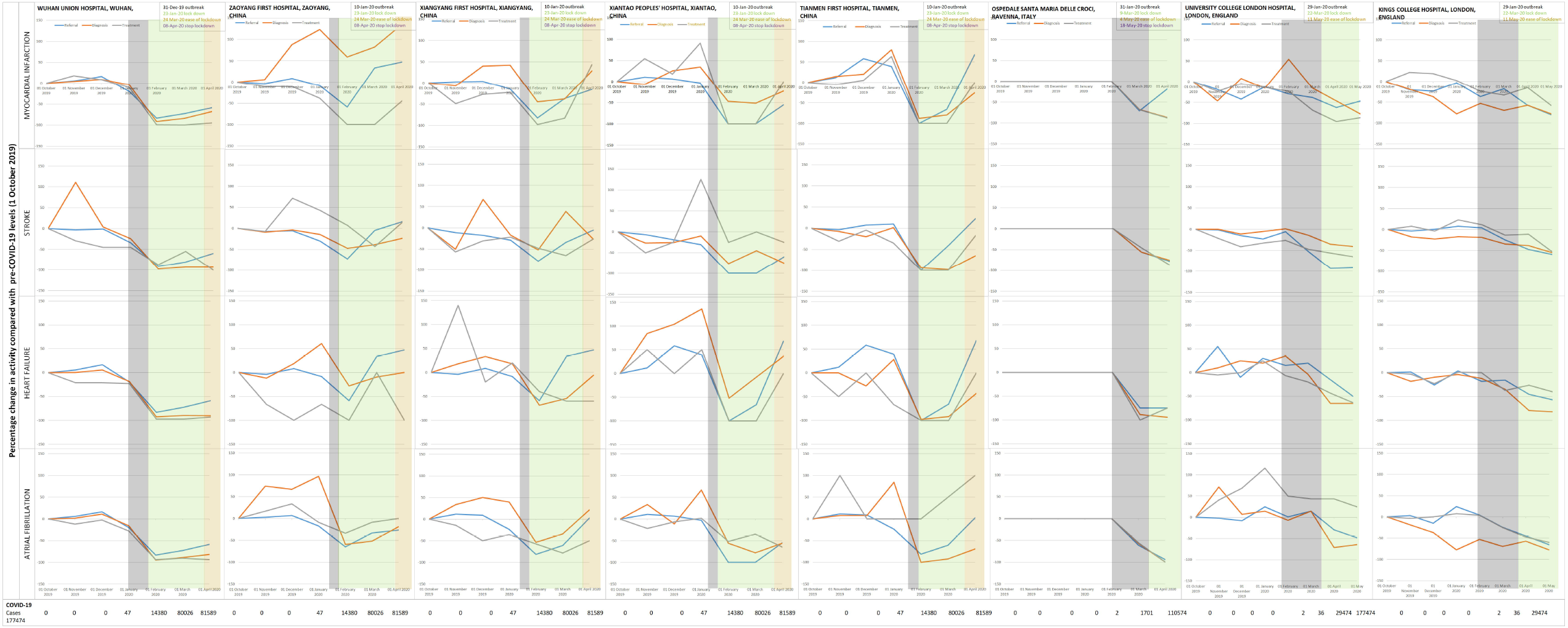

